# Parasitic manipulation or side effects? The effects of past *Toxoplasma* and *Borrelia* infections on human personality and cognitive performance are not mediated by impaired health

**DOI:** 10.1101/2023.01.07.23284298

**Authors:** Jaroslav Flegr, Jana Hlaváčová, Jan Toman

## Abstract

Bacteria *Borrelia burgdorferi s. l*. and even more the protozoan *Toxoplasma gondii* are known to affect the behavior of their animal and human hosts. Both pathogens infect a significant fraction of human population, and both survive in the host’s body for a long time. The resulting latent infections used to be considered clinically asymptomatic. In the last decade, however, numerous studies have shown that this view may be wrong and both infections can have various adverse effects on human health. Their specific behavioral effects may thus be merely side effects of the general impairment of patients’ health. We tested this hypothesis on a cohort of 7,762 members of internet population using a two-hour-long survey consisting of a panel of questionnaires and performance tests. Our findings confirmed that subjects infected with *Toxoplasma* were in worse physical and mental health and those infected with *Borrelia* were in worse physical health than corresponding controls. The infected and noninfected subjects also differed in several personality traits (conscientiousness, pathogen disgust, injury disgust, Machiavellianism, narcissism, tribalism, anti-authoritarianism, intelligence, reaction time, and precision). Majority of the behavioral effects associated with *Borrelia* infection were the same as those associated with *Toxoplasma* infection, but some dramatically differed (e.g., performance in the Stroop test). Path analyses and nonparametric partial Kendall correlation tests showed that these effects were not mediated by impaired health of the infected individuals. The results thus contradict predictions of the side effects hypothesis.

## 1. Introduction

About 20–30% of Czech population have anti-*Toxoplasma* or anti-*Borrelia* IgG antibodies in their sera. Based on histological data and existing knowledge regarding reactivation of toxoplasmosis in immunocompromised individuals, e.g., AIDS patients (Luft and Remington, 1992), one can assume that *Toxoplasma*-seropositive subjects carry dormant but viable parasites in tissue cysts in various organs of their bodies until the end of life. The presence of anti-*Toxoplasma* IgG antibodies in the serum is therefore viewed as evidence of either latent (asymptomatic) or, much more rarely, chronic (symptomatic) toxoplasmosis (Robert-Gangneux and Darde, 2012). The situation is less clear regarding *Borrelia*. It is known, however, that in this case, too, IgG seropositivity usually persists for about 10–20 years (Kalish et al., 2001). It has also been demonstrated that viable *Borrelia* spirochetes can survive for decades in biofilms in human liver, heart, kidney, and brain even after extensive antibiotic treatment (Sapi et al., 2019). This suggests that latent or chronic borreliosis might also persist for many years after the end of the acute active phase of infection.

It has long been known that *Toxoplasma*-seropositive individuals have an increased risk of many psychiatric disorders, especially schizophrenia (Torrey et al., 2012; Sutterland et al., 2015), and that they score differently in various personality (Flegr and Hrdý, 1994; Flegr and Horáček, 2017b) and performance (Havlíček et al., 2001) tests than seronegative controls. Analogous behavioral changes can be induced by laboratory infection of rats and mice, which suggests that they are the effect rather than the cause of the infection (Hodková et al., 2007). In humans, differences between *Toxoplasma*-infected and *Toxoplasma*-free subjects usually increase with time elapsed since infection and the intensity of changes negatively correlates with the level of anti-*Toxoplasma* IgG antibodies (Flegr and Havlíček, 1999). This suggests that the observed differences between seropositive and seronegative individuals are due to a slow cumulation of changes that take place over years of latent infection rather than due to the (decreasing) impact of the short phase of acute infection.

Some of the changes observed in animals are relatively specific, such as for instance the change of the innate fear of cat smell to attraction to this smell in infected mice and rats (Berdoy et al., 1995; Vyas et al., 2007). Some even seem to increase the chance of transmitting the parasite from the infected host to a noninfected individual by predation by, for instance, prolongation of reaction times (Hrda et al., 2000) in infected rodents and primates (Poirotte et al., 2016), including humans (Flegr et al., 2011; 2018). The adaptiveness of changes to the host phenotype in parasites with complex life cycles is notoriously hard to prove (Cezilly et al., 2010) but it has been demonstrated in a number of cases, including the well-known fish-manipulating trematode *Euhaplorchis californiensis* (Shaw et al., 2009) or the ant-manipulating trematode *Dicrocoelium dendriticum* (Carney, 1969). The observed behavioral changes in *Toxoplasma*-seropositive individuals are thus usually also interpreted as the product of the parasite’s adaptive manipulative activity (Moore, 2002).

Much less is known about the behavioral effects of anti-*Borrelia* IgG seropositivity. In contrast to the situation with *Toxoplasma*, any association of *Borrelia* seropositivity with psychiatric disorders is most likely weak or nonexistent (Grabe et al., 2008; Hernandez-Ruiz et al., 2020; Tetens et al., 2021) but cf. (Bransfield, 2018). Some behavioral changes have been observed, but they seem to be merely transient side effects of either current or recent Lyme disease (Makara-Studzinska et al., 2017; Hundersen et al., 2021; Tetens et al., 2021).

From a clinical point of view, a life-long latent *Toxoplasma* infection or *Borrelia* IgG seropositivity is usually considered insignificant and even asymptomatic. Nevertheless, studies performed over the past decade have shown that both *Toxoplasma* and *Borrelia* IgG seropositive subjects are in worse health and have a higher incidence of many diseases than seronegative subjects do (Flegr et al., 2014; Flegr and Escudero, 2016). The most parsimonious and often suggested explanation of the behavioral changes observed in people with latent infections is thus that these changes are merely the side effects of their impaired health. The present study explores the differences in cognitive performance and personality between subjects seropositive and seronegative for *Toxoplasma* and *Borrelia*. It focuses on testing the side effects hypothesis, i.e., it explores whether the differences in personality and cognitive performance disappear when the effect of the seropositive subjects’ worse physical and psychical health is controlled for.

## 2. Materials and Methods

### 2.1. Participants

An electronic survey consisting of several questionnaires and performance tests, with only some related to the present study, was advertised on Facebook and Twitter as a project “studying interconnections between moral attitudes, cognitive performance, and various biological, psychological, and sociodemographic factors.” At the beginning of the first questionnaire, participants were informed that the study is anonymous, and they can terminate their participation at any time. They were also provided with the following information: “We will investigate which biological and psychological traits influence your performance test scores and moral attitudes. We will measure your memory, speed, ability to concentrate, and intelligence.” Only subjects who confirmed that they were older than 15 and provided informed consent by pressing the corresponding button were allowed to participate in the study. The study was at least partially completed by 8,800 subjects between March and June 2022. The project, including the method of obtaining informed consent, was approved by the Institutional Review Board of the Faculty of Science, Charles University (No. 2021/4).

### 2.2. Questionnaires and tests

In the survey, we measured the *intelligence* of subjects with the Cattel 16PF test (variant A, scale B) (Cattell and Mead, 2008) and their *memory* with a modified Meili test (Meili, 1961; Flegr et al., 2012). In the latter test, participants were at the beginning shown 12 words (knife, frog, pump, chain, tree, collar, ice, glasses, arrow, train, bars, rifle) for 24 seconds and then, the middle of the survey (about 20 minutes into it), they were asked to identify these words from a list of 24 words. Psychomotor performance (*reaction time* and *precision* of reaction) was measured using the Stroop test. This variant of the Stroop test had three parts separated by time needed to read new instructions and to take some rest. In part A, probands had to select a specified word (for instance “red”) with their pointing device from a set of four words (“red,” “green,” “blue,” “brown”) displayed in a random order in the central part of the screen. The four words were written in red, green, blue, or brown font, and the meaning of the words did not match the font color. The command specifying which word they were supposed to select was written in the upper part of the screen. The probands were instructed to ignore the font color. In part B, the stimuli were the same but the probands were asked to select a word written in a particular color and ignore the meaning of the displayed words. Part C was similar to part A but the command specifying which word the proband should select was always written in a different color and that color matched neither the meaning of the word nor the color of the displayed stimuli. At the beginning of each part, probands received instructions about the rules of the following subtest, informed about how many times the test would run (always five times), and were asked to react as quickly as possible. The probands could start each part of the Stroop test by pressing a “Start test” button.

Five personality traits, *extroversion, agreeableness, conscientiousness, emotional stability*, and *openness to new experiences*, were measured using the Ten Items Personality Inventory (Gosling et al., 2003). Four facets of disgust (*pathogen disgust, sexual disgust, moral disgust*, and *injury disgust*) were measured with the Czech version of the Three Domain Disgust Scale (TDDS) (Tybur, 2009) supplemented with the *injury disgust* scale (Kupfer and Le, 2018). Three aspects of the dark triad (*Machiavellianism, narcissism*, and *psychopathy*) were measured using the Czech version of the Short Dark Triad test (SD3) (Jones and Paulhus, 2014; Mejzlíková et al., 2018) and four aspects of political attitudes (*tribalism, economic egalitarianism, cultural liberalism*, and *anti-authoritarianism*) were measured using the Political Beliefs and Values Inventory (PI34) (Kopecky et al., 2022).

In the anamnestic part of the questionnaire, participants responded to 11 questions concerning their physical health, namely about the frequency of suffering from infectious diseases, headaches, other physical pains, other chronic or recurring physical problems, frequency of visits to physicians, frequency of feeling tired, neurological diseases, the number of drugs prescribed by a physician (not for mental health) they were currently using, how many times they used antibiotics in the past year, how many times they used antibiotics in the past three years, how many times they spent more than a week in a hospital in the past five years, and how long they expected to live. Along similar lines, they were asked four questions concerning their mental health (frequency of suffering from depression, anxiety, other mental health problems, and how many kinds of drugs prescribed by a physician for mental health problems they were currently using). Participants answered these questions using 6-item Likert scales, the sole exception being the two questions about the number of kinds of drugs they were using; for details, see (Flegr et al., 2021). We computed indices of physical and mental sickness as the mean Z-score of the corresponding eleven or four questions, respectively. In this part of the survey, respondents were also asked about their age, sex (males coded as 1, women coded as 0) and whether they were infected with *Toxoplasma* and/or *Borrelia* (1: “I do not know, I am not sure, I have not been tested,” 2: “No, I was tested and the result was negative,” 3: “Yes, I was tested and the result was positive”). For both toxoplasmosis and borreliosis, the questionnaire was preset to indicate as a default the first response (“I do not know, I am not sure, I have not been tested”). Infected individuals were coded 3, noninfected ones 2.

### 2.3. Data analyses

The effects of toxoplasmosis and borreliosis were tested with a nonparametric partial Kendall test controlled either for sex and age or for sex, age, physical health, and mental health (Flegr and Flegr, 2021). The effect of multiple tests was controlled with the Benjamini-Hochberg procedure (FDR = 0.10). The results of the nonparametric analysis were confirmed with a parametric path analysis using Jamovi 2.2.5 (Jamovi, 2021), PATHj module (Galuci, 2021) and yEd Graph Editor (yWorks GmbH, https://www.yworks.com/products/yed, accessed 5. 11. 2022). The dataset is available at Figshare (Flegr, 2022b).

#### Technical notes

Except for the section Discussion, we use “toxoplasmosis” (“borreliosis”) or *Toxoplasma*-(*Borrelia*-) infection as an abbreviation for “reported past anti-*Toxoplasma* (*Borrelia*) seropositivity.” Moreover, throughout the manuscript we use the term “effect” in a sense common in statistics, that is, to mean an observed association. The sole place where we discriminate between cause and effect is the Discussion section. In the result section, “p << 0.001” means p < 0.000001.

## 3. Results

The raw dataset contained 4,942 women (mean age = 43.16, sd = 12.52) and 2,820 men (mean age = 39.80, sd = 12.42). Toxoplasmosis status (codes: negative = 0, positive = 1) was reported by 796 women (23.9% were positive) and 166 men (12.0% were positive). The effect of sex on *Toxoplasma* seropositivity was significant (OR = 0.437, C.I._95_ = 0.252-0.724, p < 0.001). *Toxoplasma*-infected women were older (45.0, sd = 10.16) than *Toxoplasma*-free women (42.2, sd = 11.5), p = 0.001, Cohen d = 0.26. Likewise, *Toxoplasma*-infected men were older (42.7, sd = 12.5) than *Toxoplasma*-free men (39.5, sd = 10.3), p = 0.218, Cohen d = 0.26. Borreliosis status (codes: negative = 0, positive = 1) was reported by 1,247 women (519, i.e., 41.6% were positive) and 531 men (163, i.e., 30.70% were positive). The effect of sex on *Borrelia* seropositivity was highly significant (OR = 0.621, C.I._95_ = 0.497-0.775, p << 0.001). *Borrelia*-infected women were older (45.8, sd = 12.7) than *Borrelia*-free women (42.8, sd = 12.8), p = 0.001, Cohen d = 0.32. *Borrelia*-infected men were also older (44.4, sd = 12.8) than *Borrelia*-free men (37.9, sd = 12.8), p<< 0.001, Cohen d = 0.51. We found a positive association between *Toxoplasma* and *Borrelia* positivity (OR = 3.34, C.I._95_ = 2.11-5.28, p << 0.001).

Table 1 shows the mean scores for variables related to performance and personality for all subjects as well as separately for women, men, *Toxoplasma*-negative subjects, *Toxoplasma*-positive subjects, *Borrelia*-negative subjects, and *Borrelia*-positive subjects. Many behavioral variables had an irregular distribution. Therefore, we used nonparametric multivariate tests, namely partial Kendall correlation tests, to search for associations between *Toxoplasma* or *Borrelia* seropositivity and health, cognitive performance, and personality of the participants (Table 2, left part). The results showed that *Toxoplasma*-seropositive subjects were in worse physical and mental health and *Borrelia*-seropositive subjects were in worse physical health than the corresponding controls. The tests also revealed several significant associations between the infections and behavioral variables, whereby most of these associations remained significant even after correction for multiple tests. The right part of Table 2 shows the results of analogical analyses performed with partial Kendall correlation tests controlled not only for sex and age but also for physical and mental health. A comparison of the left and right parts of the table shows only very small differences. This suggests that poor physical or mental health does not play a mediating role in the association between infections and behavioral variables. The same analyses were performed separately for women and men (Table 3).

**Table 1.**
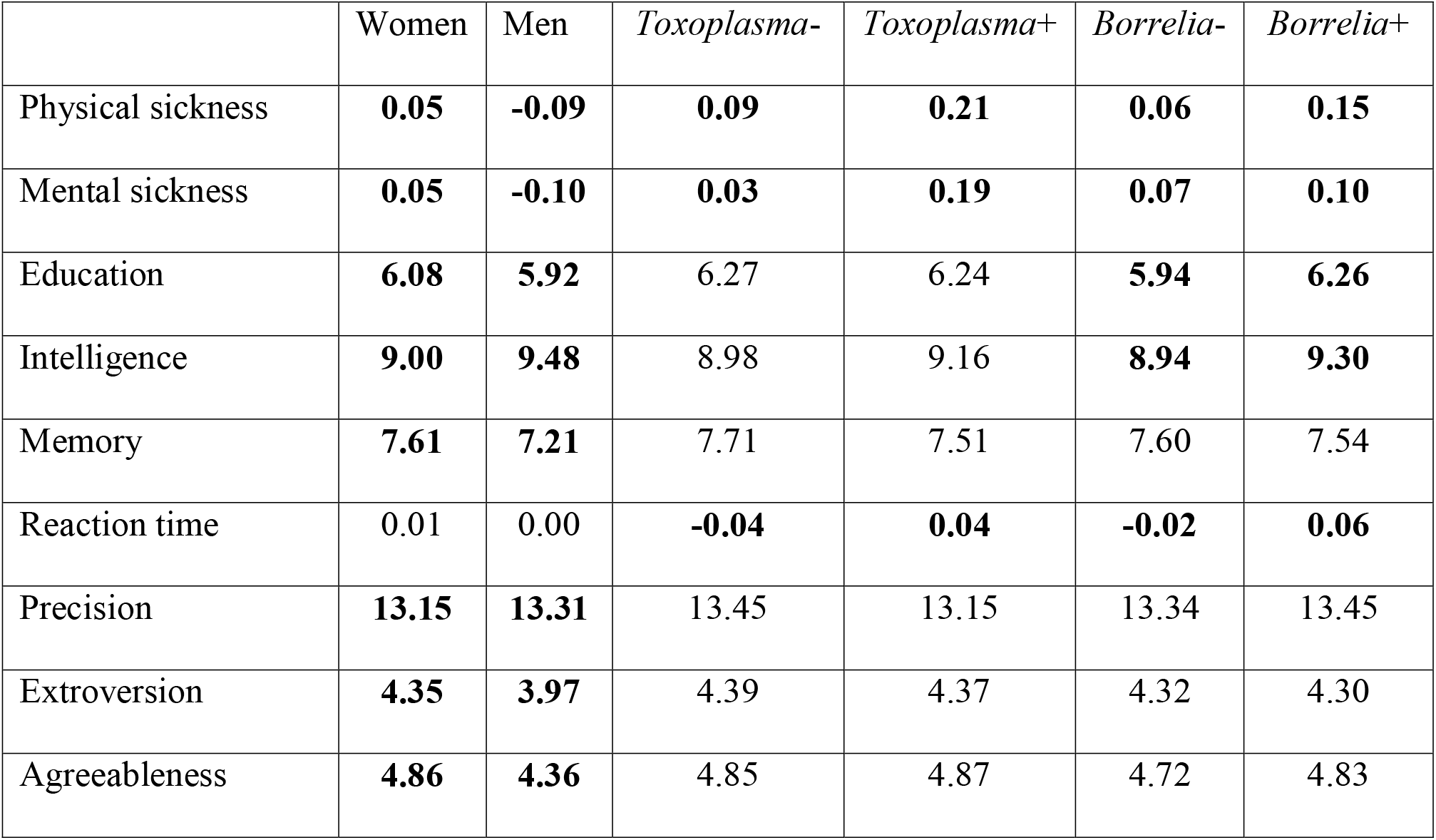
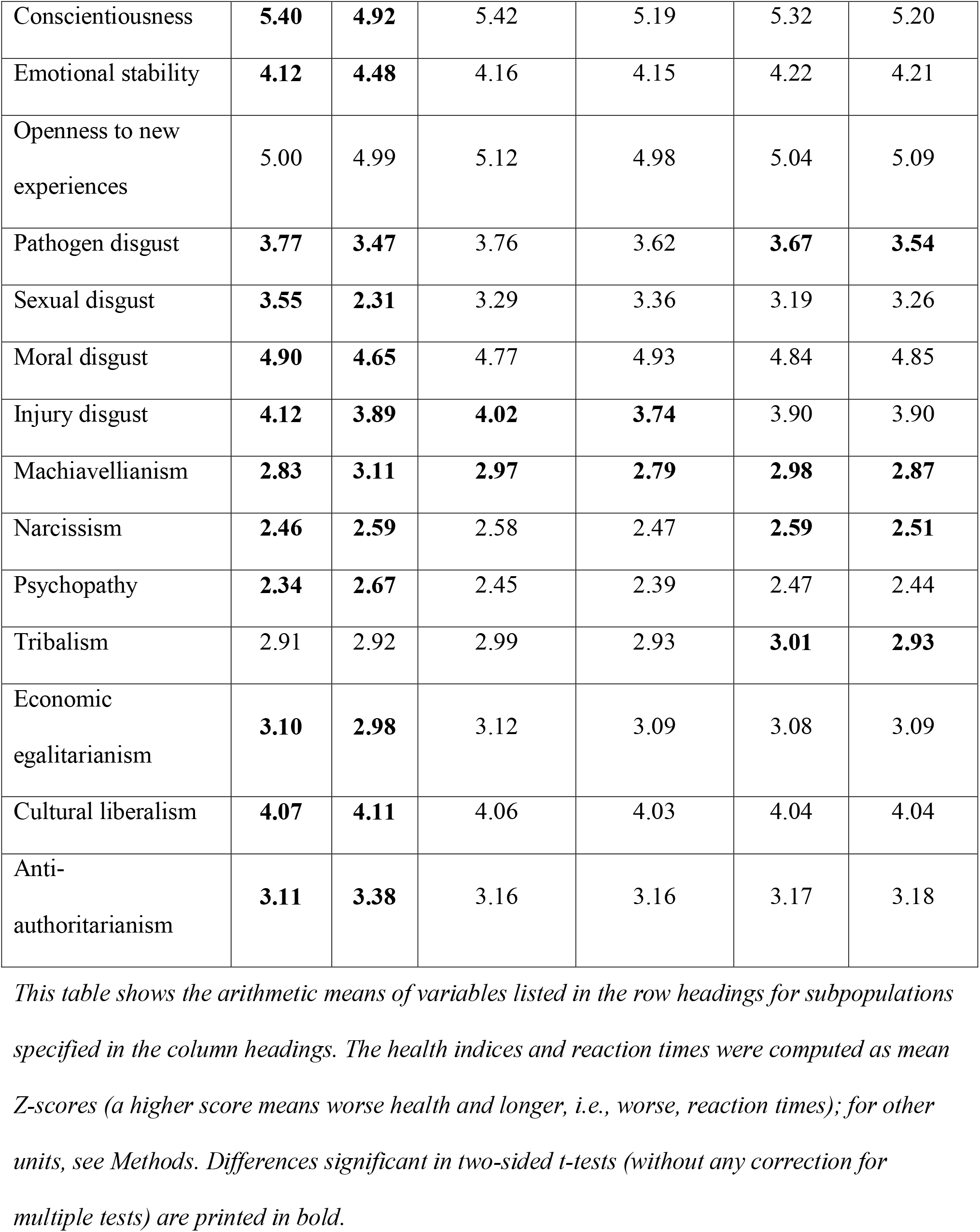
Differences in health and behavioral variables between female and male, *Toxoplasma* seronegative and seropositive, and *Borrelia* seronegative and seropositive participants.

**Table 2.** Association of infections with physical and mental health and various behavioral traits.

|  | Controlled for sex and age |  | Controlled for sex, age, and health |  |
| --- | --- | --- | --- | --- |
|  | Toxoplasmosis | Borreliosis | Toxoplasmosis | Borreliosis |
| Physical sickness | **0.058\*** | **0.049\*** | 0.042 | **0.052\*** |
| Mental sickness | **0.055\*** | 0.000 | 0.038 | -0.018 |
| Education | -0.001 | **0.049\*** | 0.005 | **0.052\*** |
| Intelligence | **0.066\*** | **0.111\*** | **0.071\*** | **0.113\*** |
| Memory | -0.011 | 0.015 | -0.006 | 0.019 |
| Reaction time | 0.038 | 0.010 | 0.036 | 0.009 |
| Precision | -0.024 | **0.037\*** | -0.022 | **0.036\*** |
| Extroversion | -0.016 | -0.023 | -0.010 | -0.020 |
| Agreeableness | -0.003 | 0.016 | 0.002 | 0.018 |
| Conscientiousness | **-0.091\*** | **-0.066\*** | **-0.085\*** | **-0.065\*** |
| Emotional stability | -0.003 | 0.002 | 0.018 | 0.006 |
| Openness to new experiences | **-0.047** | 0.018 | -0.044 | 0.020 |
| Pathogen disgust | **-0.048** | **-0.049\*** | **-0.051** | **-0.052\*** |
| Sexual disgust | -0.024 | -0.032 | -0.024 | -0.033 |
| Moral disgust | 0.009 | -0.015 | 0.012 | -0.015 |
| Injury disgust | **-0.056\*** | 0.002 | **-0.056\*** | 0.001 |
| Machiavellianism | **-0.072\*** | -0.030 | **-0.077\*** | -0.031 |
| Narcissism | -0.044 | **-0.039\*** | -0.043 | **-0.037\*** |
| Psychopathy | -0.007 | 0.028 | -0.014 | 0.027 |
| Tribalism | **-0.047** | **-0.065\*** | **-0.049** | **-0.068\*** |
| Economic<br>egalitarianism | -0.026 | 0.002 | -0.033 | 0.000 |
| Cultural liberalism | -0.006 | 0.024 | -0.005 | 0.024 |
| Anti-authoritarianism | 0.017 | **0.038\*** | 0.019 | **0.038\*** |

The first two columns of the table show the effects (partial Kendall $\tau$ ) controlled only for sex and age, while columns 3 and 4 show the results controlled for sex, age, physical health, and mental health. (The effects of infections on mental health were controlled only for sex, age, and physical health, and the effects of infections on physical health were controlled only for sex, age, and mental health). Negative $\tau$ indicates a negative association with the infection, while positive $\tau$ indicated positive associations. Significant effects are printed in bold, and asterisks indicate the effects significant after the correction for multiple tests.

**Table 3.**
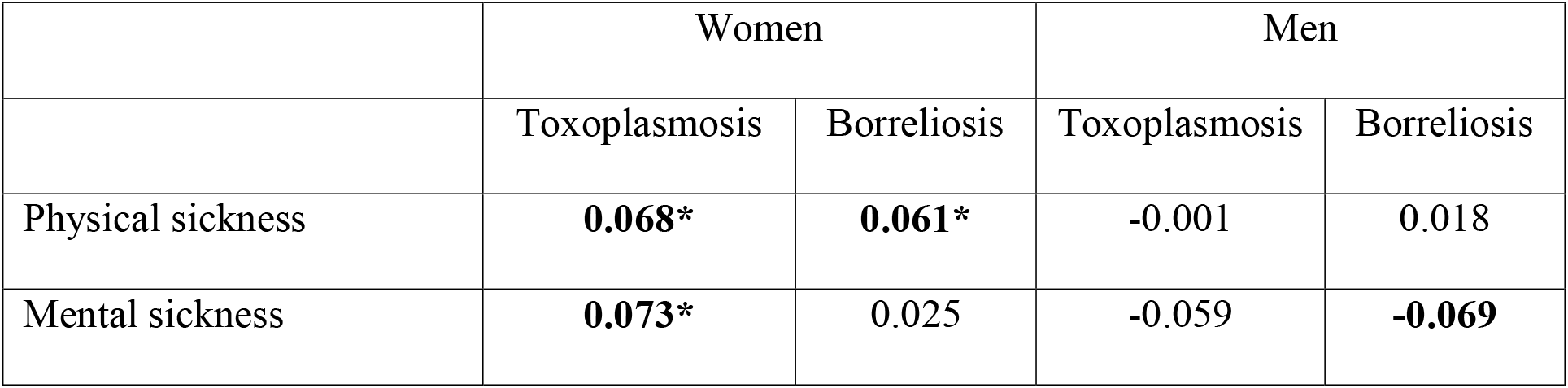
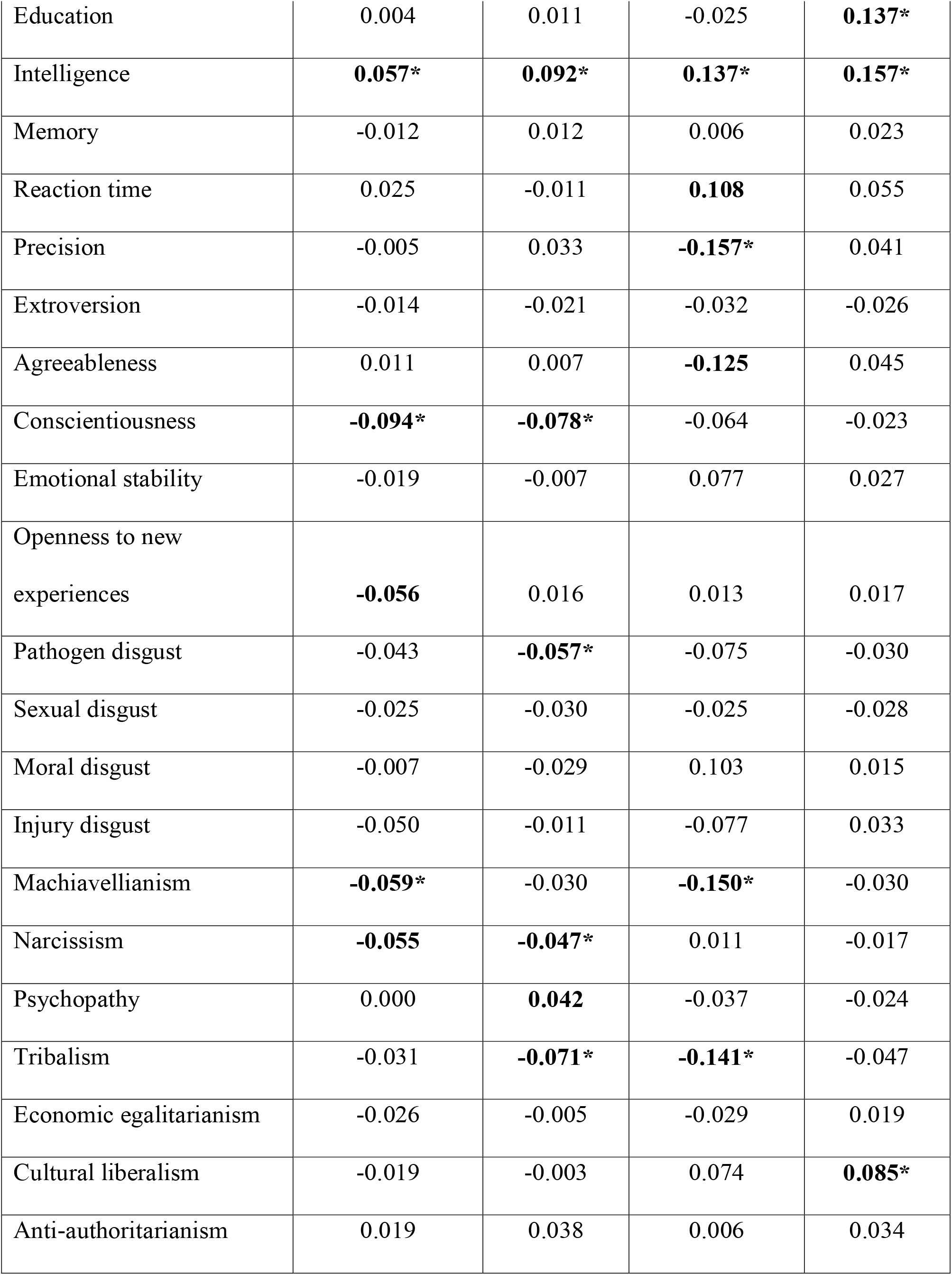
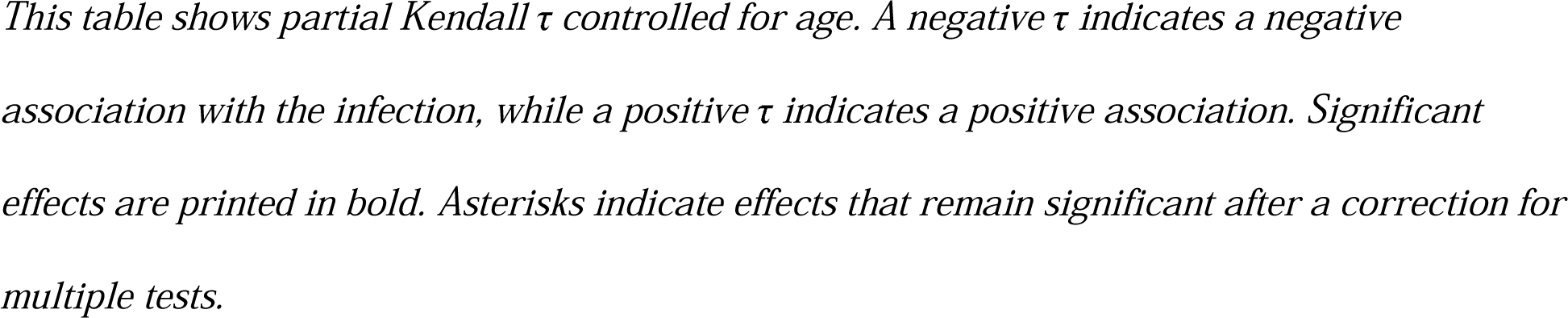
Association of infections with physical and mental health and various behavioral traits in women and men.

We used a structural equation modeling approach to explore the relationship between infections, health, and behavioral variables in more detail. Using path analysis, we confirmed the results of (nonparametric) partial Kendall correlation tests for all examined behavioral variables significantly associated with the infections in partial Kendall tests. The direct effect of infections either remained significant or the effects of impaired health and the infection on a behavioral trait went in opposite directions (Fig. 1, 2). This means that neither the more robust nonparametric nor the more sensitive parametric methods support the existence of a mediating effect of impaired health.

**Fig. 1.**
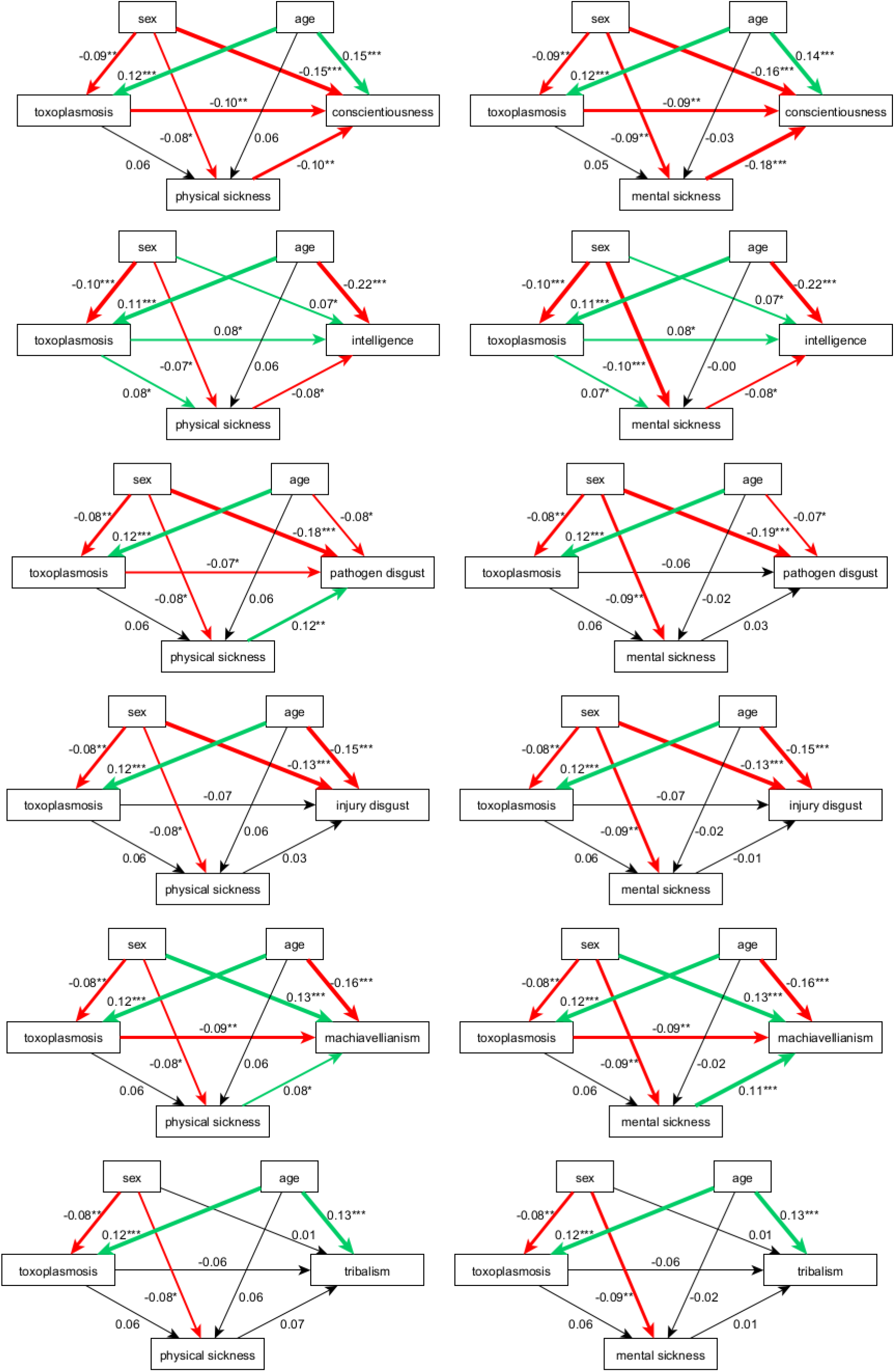
The results of path analyses for toxoplasmosis: investigation of possible mediating effects of physical or mental health. *Green arrows show positive, red arrows negative, and black arrows nonsignificant correlations. The numbers (standardized path coefficients) and arrow widths indicate the strength of correlations. The number of asterisks (one, two, or three) indicates their significance (0.05, 0.01, and 0.001, respectively). Men are coded with 1, women with 0. A negative path coefficient indicates that, for example, men scored lower on conscientiousness than women did*.

**Fig. 2.**
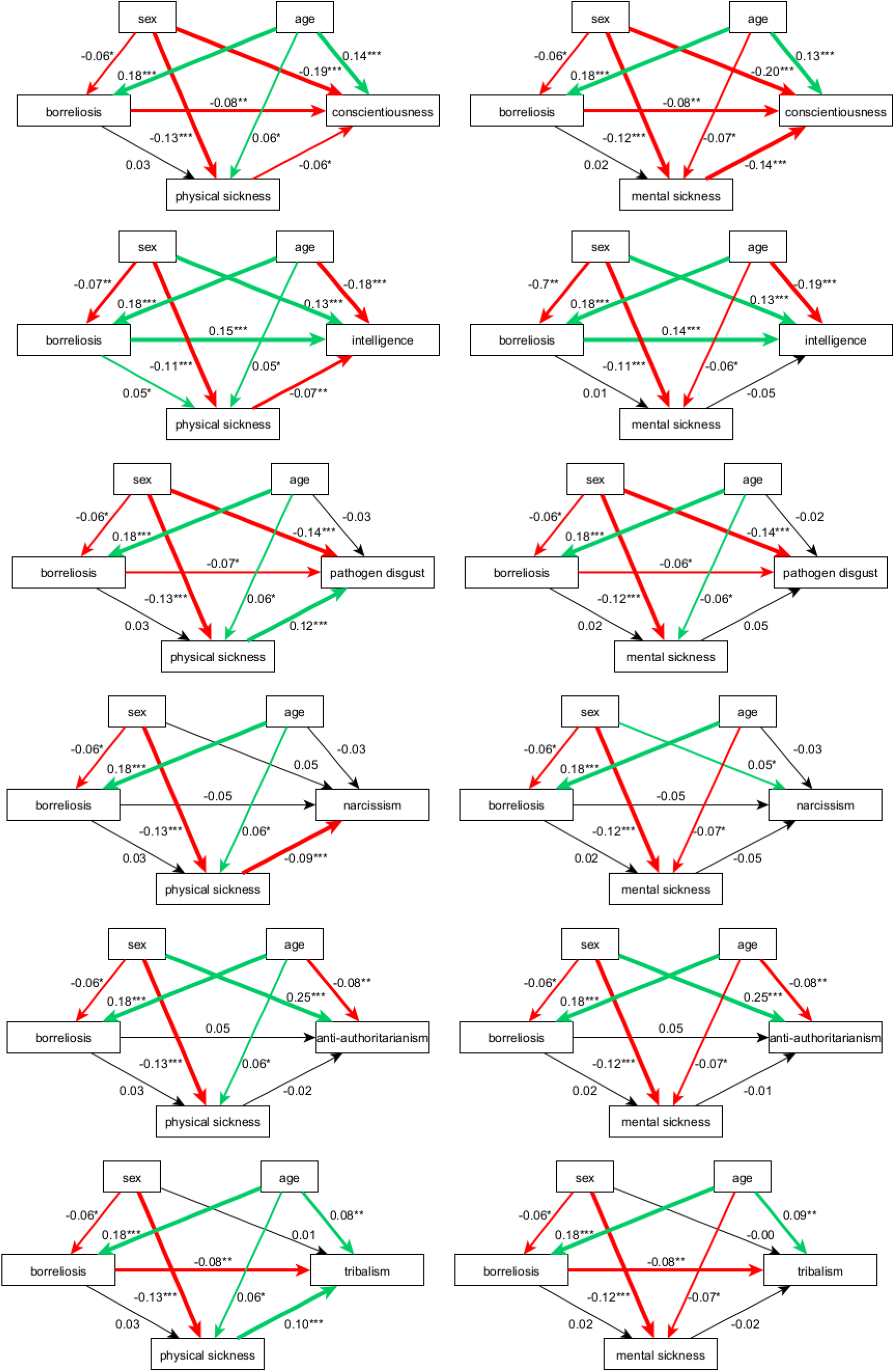
The results of path analyses for borreliosis: investigation of possible mediating effects of physical or mental health. *For legend, see Fig. 1*.

One rather counterintuitive result of the present as well as several previously published studies (Flegr and Havlíček, 1999; Flegr et al., 2013) is the positive effect of seropositivity on intelligence. We have therefore tested the hypothesis that the effect is mediated by other variables (e.g., conscientiousness), which could be affected not only by the infection but also for instance by the age or health status of the subject. Figure 3 shows the results of the corresponding path analyses. The models contained six variables, namely the participants’ age, sex, infection, physical or mental health, conscientiousness, and intelligence. The results differed for borreliosis and toxoplasmosis.

**Fig. 3.**
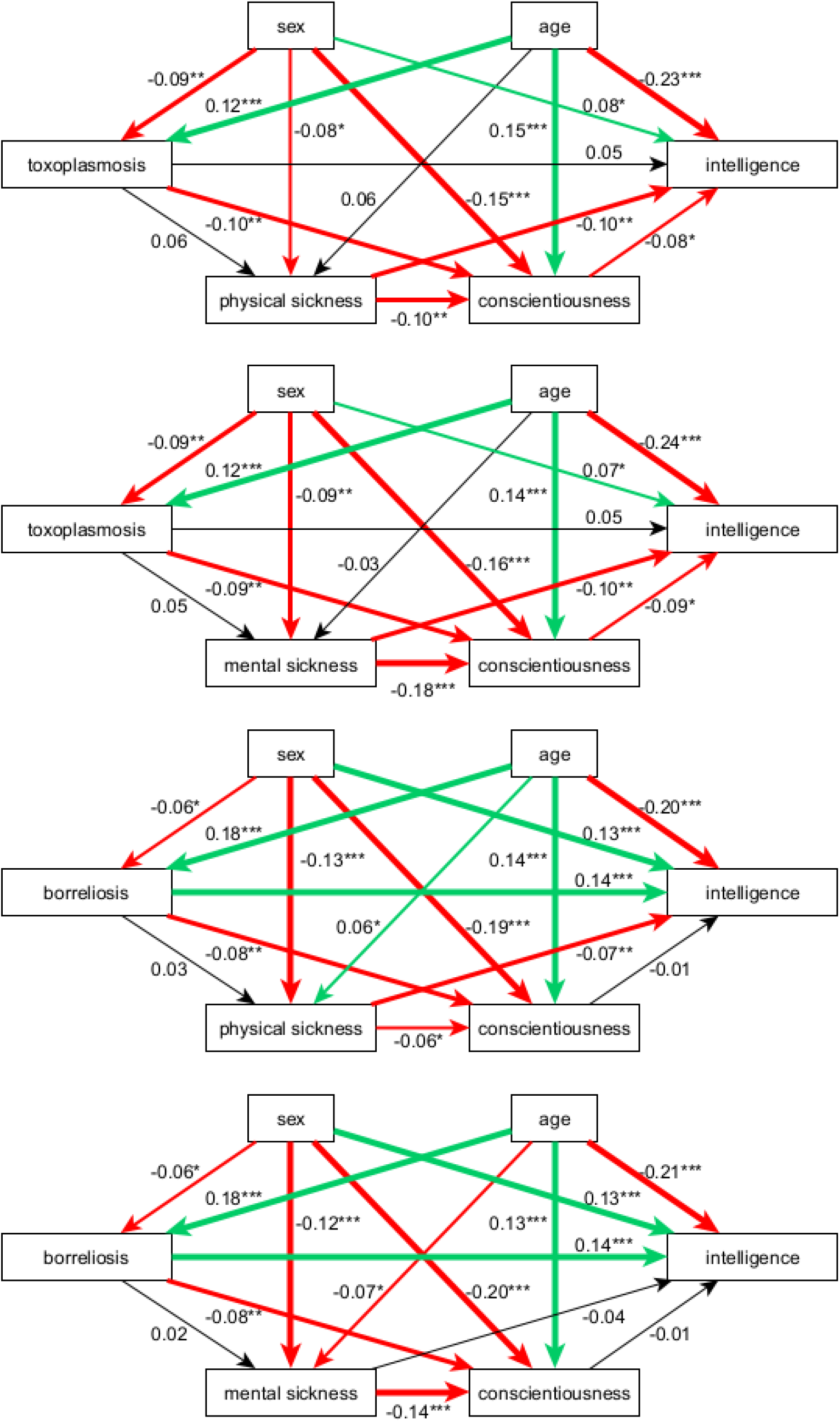
Path analysis: search for a mediating effect of conscientiousness on intelligence. *For legend, see Fig. 1*.

For toxoplasmosis, the results might partly support a model based on the mediating role of conscientiousness. When both conscientiousness and mental or physical sickness were included in the model, the direct effect (i.e., the path coefficient) decreased from 0.08 to 0.05, and thus turned formally nonsignificant. Therefore, the results suggest that toxoplasmosis negatively affects conscientiousness, conscientiousness negatively affects intelligence test results, and these two negative relations could result in a positive relationship between toxoplasmosis and intelligence. However, the strength of this indirect effect is probably weaker than the direct effect of toxoplasmosis on intelligence. Also, toxoplasmosis increased mental sickness, and mental sickness again led to decreased conscientiousness. Nevertheless, this conscientiousness-mediated positive effect of mental sickness on intelligence was canceled by a much stronger direct negative effect of mental sickness on intelligence (Fig. 3).

In the case of borreliosis, a path analysis yielded no support for a model based on the mediating effect of conscientiousness. Here, the direct positive effect of the infection on intelligence did not decrease when conscientiousness and physical or mental health were included in the models (Fig. 3, bottom part).

We performed a similar analysis to search for a possible mediation effect of education. That analysis showed a robust positive association between education and intelligence (path coefficient > 0.4; results are not shown). However, we found no association between education and *Toxoplasma* or *Borrelia* seropositivity. Therefore, education cannot be responsible for the positive association between infections and intelligence.

## 4. Discussion

In the study’s confirmatory part, we tested the hypothesis that the observed differences in cognitive performance and personality traits between anti-*Toxoplasma* or anti-*Borrelia* IgG seronegative and seropositive subjects are merely the side effects of impaired physical or mental health of the infected subjects. We found that seropositive subjects were indeed in worse health, but the results of multivariate nonparametric tests and path analyses contradicted inferences derived from the side effects hypothesis. In particular, the strength of all associations between seropositivity and behavioral traits observed in our relatively large data set remained approximately the same regardless of whether physical and mental health were controlled for or not. Moreover, for several output variables the effects of seropositivity and the effect of impaired health on behavioral variables went in opposite directions.

In the study’s exploratory part, we confirmed the existence of some previously described effects, found some yet unknown effects, and obtained some results which disagree with previously published data. In the case of toxoplasmosis, we confirmed that the infected individuals were in worse physical and mental health than the corresponding controls. This observation corresponds to results obtained using similar questionnaire-based studies as well as with results obtained using other empirical approaches, such as cross-sectional studies performed on serologically tested volunteers (Flegr and Escudero, 2016) or ecological (correlative) studies conducted in 88 countries worldwide or in 29 European countries (Flegr et al., 2014). We have also confirmed lower conscientiousness but not higher extroversion of seropositive subjects. In one past study, the Big Five traits were measured using another personality questionnaire (NEO-PI-R) in a (much younger) population of university students whose *Toxoplasma* status was serologically examined immediately before the study (Lindová et al., 2012). These differences in the experimental setup could explain the partly different findings of the two studies, namely the absence of an effect of toxoplasmosis on extroversion in the present study. We confirmed the effect of *Toxoplasma*-seropositivity on psychomotor performance. The effects were nonsignificant in women but significant and relatively strong in men and they showed both longer reaction times and lower precision (higher number of errors) in *Toxoplasma*-seropositive men. *Toxoplasma*-positive participants, both men and women, scored higher on intelligence tests than *Toxoplasma*-negative subjects did. A recent metanalytic study (de Haan et al., 2021) reported a mild impairment of cognitive functions in *Toxoplasma*-positive subjects but, for some unknown reason, this study excluded all studies that used IQ questionnaires. When the intelligence of 191 women tested for toxoplasmosis during pregnancy was essayed with the Cattel 16PF test, those who were *Toxoplasma* positive scored higher than those who were *Toxoplasma* negative (Flegr and Havlíček, 1999). In another study performed on 857 military conscripts (men), authors showed that *Toxoplasma*-positive subjects scored significantly worse in the OTIS intelligence test (Otis, 1954), But in that study, intelligence negatively correlated with anti-*Toxoplasma* antibodies titer and therefore positively with the duration of infection, which suggests that the observed lower performance of *Toxoplasma*-positive subjects in IQ tests was most likely a transient effect of recent acute toxoplasmosis rather than a cumulative effect of latent toxoplasmosis. Another group of 502 male soldiers was tested with two IQ tests: the Wiener Matrizen-Test (nonverbal general intelligence test) and the OTIS verbal intelligence test (Flegr et al., 2013). In that study, the RhD-positive *Toxoplasma*-infected subjects scored lower, while RhD-negative *Toxoplasma*-infected subjects scored higher on intelligence than their *Toxoplasma*-free peers. To summarize, *Toxoplasma*-positive and *Toxoplasma*-negative subjects usually differ in their performance in intelligence tests, but the direction of the observed association varies between studies. This suggests that the observed changes in intelligence are rather a side effect of other effects of the infection or that, possibly, some unknown variable independently influences both intelligence and the probability of being seropositive.

We found no effect of *Toxoplasma* positivity on memory as measured with a modified Meili test. In contrast, several recent studies have reported impairments of some – but not all – memory functions in *Toxoplasma*-positive subjects (Mendy et al., 2015a, b; Wiener et al., 2020). Our present results concerning political attitudes sharply contrast with those published recently in another study (Kopecky et al., 2022). In the present study, we found a negative association of *Toxoplasma* positivity with tribalism (tribal conscientiousness and loyalty, exaltation of own tribe, e.g., nation, over other groups), nonsignificant in path analysis, while in a study performed on data collected by the same method earlier (2016–2018), the association between *Toxoplasma* positivity and tribalism was positive. We can only speculate about the reasons which may be driving this change. It is, for example, possible that a dramatic shift in political attitudes which took place in the Czech population after the beginning of the Russian invasion of Ukraine may have affected the nature of association between toxoplasmosis and tribalism. Before the war in Ukraine, tribalism (especially in its extreme forms) was very unpopular among altruistic participants of our (long and unpaid) questionnaire studies. The war in Ukraine, which broke out near our borders, led to an increased sense of threat by Russia and the arrival of over 300,000 Ukrainian refugees, may have changed this attitude significantly in many altruistic participants of our studies. If toxoplasmosis primarily affects altruism, then the infection might correlate negatively with tribalism before the start of the Ukraine war and positively after its start. Path analysis shows that both toxoplasma seropositivity and tribalism positively associate with age. The effect of age alone, however, does not explain the striking differences between current and previous study, As far as we know, the relationship between toxoplasmosis and the dark triad traits or disgust has not been studied before. We found that *Toxoplasma*-positive subjects, especially men, scored lower on Machiavellianism and *Toxoplasma*-positive women scored lower also on narcissism.

We included the dark triad test in the survey because we anticipated that impaired health might affect the life strategy of individuals (who would shift from a slower to a faster life strategy) (Sýkorová and Flegr, 2021), which in turn would affect at least some of these traits. Our results, however, showed that the effects of toxoplasmosis and impaired health on these traits went in the opposite direction.

We found a robust negative association of pathogen disgust and injury disgust (in this case nonsignificant but still relatively strong in path analysis) with *Toxoplasma* positivity. It is possible that higher levels of pathogen and injury disgust could protect people against *Toxoplasma* infection. It is essential to note that pathogen disgust was negatively associated with both *Toxoplasma* and *Borrelia* positivity, while injury disgust was associated only with *Toxoplasma* positivity. One can reduce the risk of *Toxoplasma* infection by avoiding the consumption of raw meat and by increasing hygienic standards. What is less clear, though, is how higher pathogen disgust could reduce the risk of *Borrelia* infection (the risk of tick bites). One possibility is that pathogen disgust correlates with disgust towards ectoparasites, a hypothesis which finds support for instance in (Lorenz et al., 2014). It is rather difficult to explain how injury disgust could decrease the risk of *Toxoplasma* infection. *Toxoplasma* can be transmitted by blood but in modern times, this transmission route is most likely rather rare. One speculation could be that a higher propensity to eat raw meat is associated with decreased disgust in general (injury disgust included), as attested by findings according to which digestion of raw meat is a powerful disgust trigger (Angyal, 1941). Another possible explanation follows from the results of path analysis. In this analysis, injury disgust showed association with age and sex that may possibly drive both the probability of becoming toxoplasma seropositive and the differences in injury disgust.

In the present study, we confirmed that *Borrelia*-positive individuals were in worse physical but not mental health than the corresponding controls (Flegr and Horáček, 2018). Behavioral effects of borreliosis are studied very rarely. The search for the combination of terms toxoplas* AND behavio* yielded 1,008 hits, about 98% of which focused on the effects of toxoplasmosis on the behavior of intermediate mammal hosts or the mental health of humans (Web of Science, Core Collection, 17. 8. 2022). In contrast, an analogical search for borrel* AND behavio* resulted in 383 hits, only 11 (2.8%) of which dealt with the effect of borreliosis on behavior or mental health. Moreover, these 11 studies investigated either neuroborreliosis or acute borreliosis.

Therefore, the part of our study concerning the behavioral effects of *Borrelia* seropositivity had an explorative character and the observed associations cannot be compared with previously published data. If nothing else, it should be noted that the positive association of intelligence with *Borrelia* positivity was nearly two times stronger than the equivalent association with *Toxoplasma* positivity (τ 0.111 vs. 0.066).

In contrast to the situation with *Toxoplasma, Borrelia*-positive subjects scored higher on achieved education (only men) as well as precision (they achieved a higher number of correctly selected targets in the Stroop test). Similar to the situation with *Toxoplasma*, we found no effect of *Borrelia* positivity on memory. *Borrelia*-positive men and women scored lower on conscientiousness, pathogen disgust, and tribalism than their *Borrelia*-negative peers. These three negative associations were also observed with *Toxoplasma* positivity. In contrast to the situation with *Toxoplasma*, the negative association of *Borrelia* positivity with Machiavellianism was insignificant in both tests, whereas the same association with narcissism was significant only in Kendall correlation test. We also found a positive association of *Borrelia* positivity but not *Toxoplasma* positivity with anti-authoritarianism. In conclusion, majority of the behavioral effects of *Borrelia* seropositivity were the same as those observed in *Toxoplasma* seropositivity. A few, however, differed and in some cases even dramatically so (namely the performance in the Stroop test).

The main limitation of the present study is that the participants reported their serostatuses themselves. We asked them to relay only on the results of laboratory tests. However, some participants might have confused pathogens, and others might have misinterpreted the test results, possibly confusing IgG and IgM seronegativity. Also, some could acquire the infection after past negative tests. It must be reminded that such issues could result in the Type II error – the failure to detect an existing effect but not in the Type I error – detecting a nonexistent effect (Flegr and Horáček, 2017a). Another limitation of the study is that the participants were self-selected. The age structure of the sample and the general adult population of the Czech Republic was similar; however, women were strongly overrepresented in the sample. Most importantly, only altruistic and curious people probably voluntarily participated in similar (unpaid) studies. The composition of the sample, therefore, does not reflect the composition of the general Czech population, and everybody must be careful in the generalization of our results. This issue, however, concerns all studies that fulfill modern ethical standards, i.e., which explain in the informed consent the voluntary nature of participation in the study.

## 5. Conclusions

Our results suggest that the behavioral effects of latent toxoplasmosis and borreliosis are direct effects of the infections rather than side effects of impaired health of the infected subjects. It should be born in mind, however, that neither path analysis nor any type of correlation analysis can confirm the validity of a model. For example, neither path analysis nor any other statistical technique can exclude or confirm the involvement of unknown factors which are not included in the models. Moreover, a statistical analysis of observational data cannot discriminate between the cause and effect in particular associations, i.e., it cannot say anything about the direction of arrows connecting pairs of variables in the model. For example, no statistical technique can decide whether the positive association between intelligence and *Toxoplasma* or *Borrelia* seropositivity is the result of a positive (direct or indirect) effect of higher intelligence on the risk of infection or the result of a positive (direct or indirect) effect of the infection on intelligence.

Only an experimental study, not an observational study, could discriminate between these two fundamentally different models. For obvious ethical reasons, experimental studies cannot be performed on humans.

Moreover, neither observation nor experiment can decide whether a particular behavioral change is a side effect of some process associated with the infection or the product of the parasite’s manipulation activity. It is often the case that a positive effect of some behavioral change on the biological fitness of the parasite is viewed as amounting to evidence that the change is the product of manipulation activity of the parasite. It should be, however, noted that nonspecific side effects may increase the biological fitness of a parasite as well (Flegr, 2022a).

On the other hand, statistical analyses of data from observational and experimental studies can refute or at least fundamentally challenge the validity of some models. This is the case of our analysis, which in effect refuted the possibility that the observed differences in cognitive performance and personality traits are merely side effects of the impaired health of infected individuals. This model is just one of many possible models, but it was the most feasible one until the current study was completed.

## Data Availability

All data produced are available online at Figshare.

https://doi.org/10.6084/m9.figshare.21151075.v1

## Acknowledgments

We would like to thank Anna Pilátová for the final revisions of our text.

## Funding

This research was supported by Czech Science Foundation, grant number 22-20785S. Our sponsor had no involvement in the study design, the collection, analysis and interpretation of data, the writing of the report, or in the decision to submit the article for publication.

## Declarations of interest

none

## Notes

### Competing Interest Statement

The authors have declared no competing interest.

### Author Declarations

Institutional Review Board of the Faculty of Science, Charles University gave ethical approval for this work.

